# MRI-derived estimation of biological aging in patients with affective disorders in a 9-year follow-up - a prospective marker of future recurrence

**DOI:** 10.1101/2025.04.28.25326539

**Authors:** Katharina Förster, Nils R. Winter, Jan Ernsting, Ramona Leenings, Lukas Fisch, Carlotta Barkhau, Maximilian Konowski, Daniel Emden, Anna Kraus, Katharina Dohm, Klaus Berger, Volker Arolt, Angela Carballedo, Danutia Lisiecka, Thomas Frodl, Philipp Kanske, Udo Dannlowski, Tim Hahn, Dominik Grotegerd

## Abstract

**Background:** We investigated associations of the brain age gap (BAG), the difference between actual and estimated age derived from MRI scans, with disease course over nine years in patients with affective disorders in a long-term prospective design.

**Methods:** At two time-points, we acquired T_1_-weighted MRI images (mean [SD] follow-up period 8.98 [2.20] years) of patients with Major Depressive Disorder (MDD; N=32) and Bipolar Disorder (BD; N=6) and healthy controls (HC; N = 37) at two sites (Dublin, Münster). Using a brain age prediction model trained on a sample of over 10,000 subjects of the German National Cohort (GNC), we estimated individual BAG at two time-points (baseline and follow-up) using gray matter segments derived from MRI images. Employing linear-mixed-effects models, we tested main effects of diagnosis and hospitalizations during follow-up on BAG at baseline and follow-up, as well as their interaction with time respectively. In an exploratory analysis, we tested if BAG at baseline was predictive of hospitalizations during the nine-year follow-up using logistic regression and 10-fold nested cross-validation.

**Results:** MDD patients showed a larger BAG compared to HC (MDD>HC: p=.039, MDD vs. BD: n.s.), while BD patients only showed a tendency for a larger BAG (p = .066). In the Münster subsample (N=52), patients with hospitalizations showed a higher BAG compared to patients without hospitalizations (p=.001). No significant group-by-time interaction could be detected. However, higher BAG at baseline was associated with the number of hospitalizations during follow-up (p=.018), however, the cross-validation of our prediction with an accuracy of 64.3 % was not significant (p=.071).

**Discussion:** Our results show that BAG did not change over time as a function of patients’ course of disease. The present study rather suggests that a higher estimation of biological aging (higher BAG) predicts future hospitalizations. Therefore, BAG may indicate a patient’s vulnerability to future recurrence.

## Introduction

Affective disorders contribute immensely to the global burden of disease worldwide [1, 2]. While some patients do recover and are able to live with only minor health restrictions, others suffer from recurrence and report an increasing disability leading to early retirement, a reduced quality of life and elevated mortality rates [3, 4]. Consequently, affective disorders substantially contribute to years lived with disability worldwide [5].

While for many medical conditions treatment and survival rates have dramatically improved in the last decades, mortality and prevalence rates as well as standard diagnostic procedures for mental disorders have remained virtually unchanged [6]. Multiple reasons that might explain the lack of success in the discovery of neurobiological signatures and useful biomarkers have been suggested such as a lack of validity of clinical diagnoses or the heterogeneity of characteristics within patient populations with the same diagnosis, but also the primarily univariate and unidimensional statistical approach traditionally used in neuroimaging [7, 8]. To overcome these obstacles, researchers have recommended using a combination of advanced neuroimaging with machine learning methods to establish multivariate brain signatures associated with mental disorders that improve diagnostic accuracy or optimize treatment response prediction [8–10].

Recently, a novel multivariate biomarker has emerged in the field of neuroimaging, aiming to quantify the brain changes associated with aging [11]. To this end, so-called “brain age gaps” (BAG) can be calculated to estimate the “biological age” of an individual’s brain [12, 13]. First, a multivariate machine learning model is trained on a normative population to predict chronological age from structural MRI data. Second, a disease population is fed to the trained model which estimates brain age of the individual patient. Finally, BAG estimates are calculated for every patient by subtracting chronological from predicted age. Positive BAGs thus indicate an accelerated brain aging in comparison to the normative age trajectory. As the brain age approach is based on a multivariate model, it makes use of the multi-dimensionality of neuroimaging data and is therefore able to integrate all gray matter information indicative of age. This complex pattern of age-related changes is then integrated into a single biomarker. The underlying hypothesis of the brain age prediction paradigm is that this brain age gap may serve as a marker of disease risk and there are a number of studies emphasizing an association between brain age gaps and clinically relevant variables [14, 15]. For instance, a brain age model trained with gray matter segments from healthy people was predictive of a history of stroke, diabetes, and alcohol intake as well as cognitive performance in a large sample of > 14,000 individuals from the UK Biobank [16]. Other studies show an association with general mortality as well as neurological disorders such as multiple sclerosis, mild cognitive impairment (MCI) and Alzheimer’s disease [17–19]. Importantly, brain age gaps are not only associated with diagnosis per se but have been found to be predictive of the illness trajectory, predicting severity in multiple sclerosis or conversion to Alzheimer’s disease [20, 21]. Finally, brain age gaps appear to be altered not only in neurological diseases but also across mental disorders and risk factors for mental disorders, such as early life adversity [22–24]. So far, however, most studies investigate cross-sectional differences in brain age gaps between disorders. Thus, to what extent increased BAGs are a consequence or predictor of mental disorders still remains unclear, although some evidence suggests increasing BAGs after disease onset [25].

Here, we apply an independent brain age model trained on a large normative sample of over 10,000 subjects to a longitudinal sample of patients with affective disorders measured at a large time interval, averaging 9 years. Our primary goal was to investigate whether brain age estimates can serve as a prospective biomarker for the course of disease in patients with affective disorders.

1. We expect that patients with affective disorders show an increased BAG compared to HC (main effect diagnosis, diagnosis model).
2. Given that alterations in gray matter volume have previously been linked to a more adverse disease trajectory and an increased number of hospitalizations in affective disorders [26– 29], we expect patients with a recurrent disease course between scans, that is having new hospitalizations, to show higher BAGs than patients without further hospitalizations and healthy controls (main effect hospitalization, hospitalization model).
3. We expect patients with a recurrent disease course in the interval to show an increase in BAG at follow-up, while patients who recover over time will show a decrease in BAG over time (course of disease by time interaction, hospitalization model).

## Methods

### Participants and Procedure

A total of 86 participants were investigated. N = 75 individuals remained for the analysis of the data after MRI preprocessing and quality control (Dublin: N =23, Münster: N = 52). In Münster, 35 patients with affective disorders (BD = 10, MDD = 25) and healthy controls (N = 26) were investigated at follow-up between 8 and 12 years after their initial recruitment. For further information see our recent publication on longitudinal gray matter changes in the same sample [29].

### Material

#### Course of disease

Patients who reported no hospitalization between scans were categorized as “nonhospitalized” (N =15). Patients who reported a hospitalization between scans were categorized as “hospitalized” (N=13). The analysis was restricted to the Münster subsample because all patients in the Dublin sample received outpatient treatment.

### MRI acquisition and preprocessing/VBM segmentation

Information on MRI acquisition, preprocessing and Voxel-Based Morphometry (VBM) segmentation can be found in supplemental material S1.

### Brain age gap

To calculate subject-specific brain age gaps, we used a previously developed brain age prediction model that was trained on a large representative sample of over 10,000 individuals of the German National Cohort (GNC). The brain age model is based on a Monte-Carlo Dropout Composite-Quantile-Regression Neural Network (MCCQR-NN). For more details regarding the model architecture and training procedure, see [30]. In short, in contrast to existing brain age models, the MCCQR-NN model provides accurate estimations of predictive uncertainty in high-dimensional neuroimaging data while ensuring state-of-the-art model performance. It is therefore especially suited for the detection of subtle brain age changes, e.g., in clinical cohorts. All brain age predictions were made using the Python machine learning package PHOTONAI [31].

Brain age gaps are defined by the deviation between an individual’s chronological age and the age prediction provided by the brain age model. First, we fed unsmoothed gray matter voxel data to the model to retrieve brain age predictions. Second, to calculate brain age gaps, we subtracted the individual’s chronological age from the predicted age. Therefore, a positive BAG indicates accelerated while a negative BAG indicates decelerated brain aging.

As a single brain age prediction can vary in its predictive certainty, the MCCQR-NN additionally provides an estimation of the epistemic and aleatory uncertainty. This uncertainty estimation can be used to calculate uncertainty-corrected BAG z-scores common in normative modeling approaches [32]. Intuitively, correcting brain age predictions for their respective uncertainty pulls highly uncertain predictions towards the population norm, therefore reducing the risk of falsely classifying individuals as deviating from the normative distribution. For all analyses, we used uncertainty-corrected z-scores as primary measure. To make the interpretation easier, we used raw BAGs (measured in years) for all figures.

### Statistical Analysis

We employed two multi-level linear-mixed effect models using the *nlme* function implemented in R [33] nested within the person controlling for age at baseline, site and length of the follow-up interval. In the first model, patients were categorized into a group according to their diagnosis (MDD or BD), further referred to as the *diagnosis model*. In the second model, limited to the Münster subsample, patients were categorized into a group according to their course of disease in the interval: patients with further hospitalizations between scans were categorized as “hospitalized patients”, while patients without further hospitalizations were categorized as “non-hospitalized patients”. We will further refer to this model as the *hospitalization model*. We then calculated between-subjects effects (main effect group and time) and a *group x time* interaction.

### Exploratory Analysis

We conducted an exploratory analysis of variance (ANOVA) with the main effect of group (patients with hospitalizations until follow-up, patients without hospitalization until follow-up and HC) on BAG at baseline only, while controlling for age and length of follow-up interval. By excluding patients with bipolar disorder and controlling for medication intake in a patient only analysis, we ensured the validity of our results in a secondary analysis (see supplemental material S2).

To follow up on our exploratory results, we conducted a logistic regression using the caret-package in R [34] predicting hospitalization during follow-up in the Münster sample using BAG (z-scores) at baseline while controlling for age at baseline, length of follow-up interval and diagnosis. We calculated predictive accuracy using 10-times repeated 10-fold cross-validation. Significance of the predictive model was assessed using a permutation test with 1000 permutations. Data and reproducible scripts can be found in the open science framework https://osf.io/qadxz/.

## Results

### 1) Diagnosis effects

The mixed linear effects model yielded a significant main effect of diagnosis (χ^2^(10) = 7.527, p = .023): MDD patients differed from HC in their BAG (MDD > HC: t(75) = 2.202, p = .039) while BD showed a tendency to differ from HC (BD > HC t(69) = 1.868, p = .066); see also *Figure 1*. BAG of BD and MDD patients did not differ (BD > MDD: t(69)=9.693, p =.490), This effect was not modulated by time (χ^2^(12) = 0.448, p = .79). We found a main effect of time (χ^2^(8) = 32.082, p < .001) and age (χ^2^(5) = 29.934, p < .001) with BAG being lower for older participants (t(69) = -6.861, p < .001) and decreasing over time (t(72) = 3.611, p < .001).

**Figure 1.**
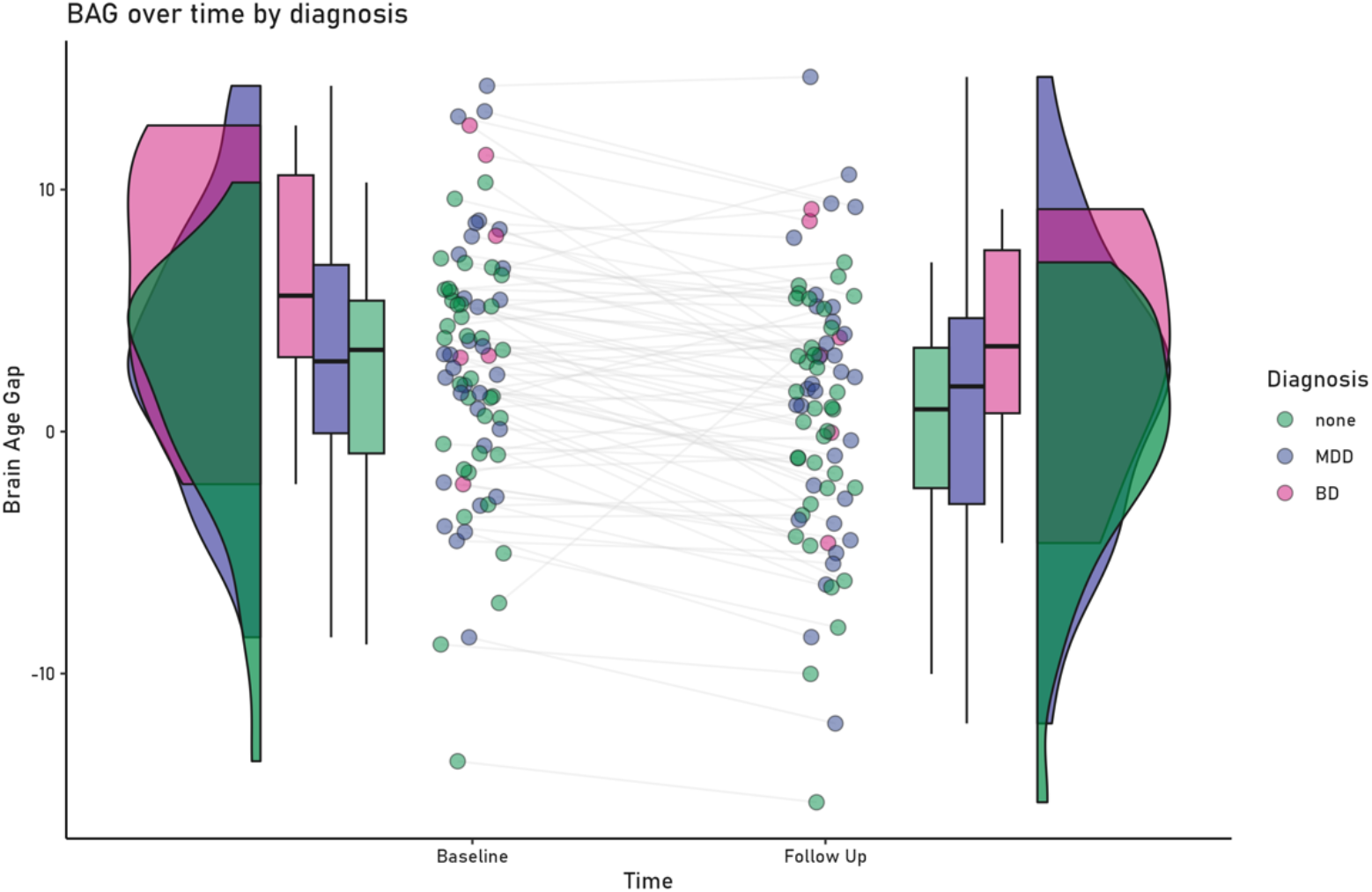
Brain age gap at baseline and follow-up for the healthy control group and patients with Major Depressive Disorder (MDD) or Bipolar Disorder (BD).

### 2) Effect of hospitalizations during the follow-up interval

The mixed-linear-effects model yielded a significant main effect of group (χ^2^(9) = 12.281, p = .002): While hospitalized patients showed a higher BAG than HC (t(47) = 3.403, p = .001) and nonhospitalized patients (t(47) = -2.2827, p = .001), nonhospitalized patients did not differ from controls (t(47)= 1.106, p = .27), see *Figure 2*. This effect was not modulated by time (χ^2^(11) = 0.519, p = .77). Including either antidepressant intake at follow-up (yes/no) and excluding BD patients from the analysis led to the same pattern of results (see *Supplement S2*). Again, there was a significant effect of age at baseline (χ^2^(5) = 14.628, p < .001) and time (χ^2^(8) = 28.621, p <.001). The direction of effect was comparable to the *diagnosis* model.

**Figure 2.**
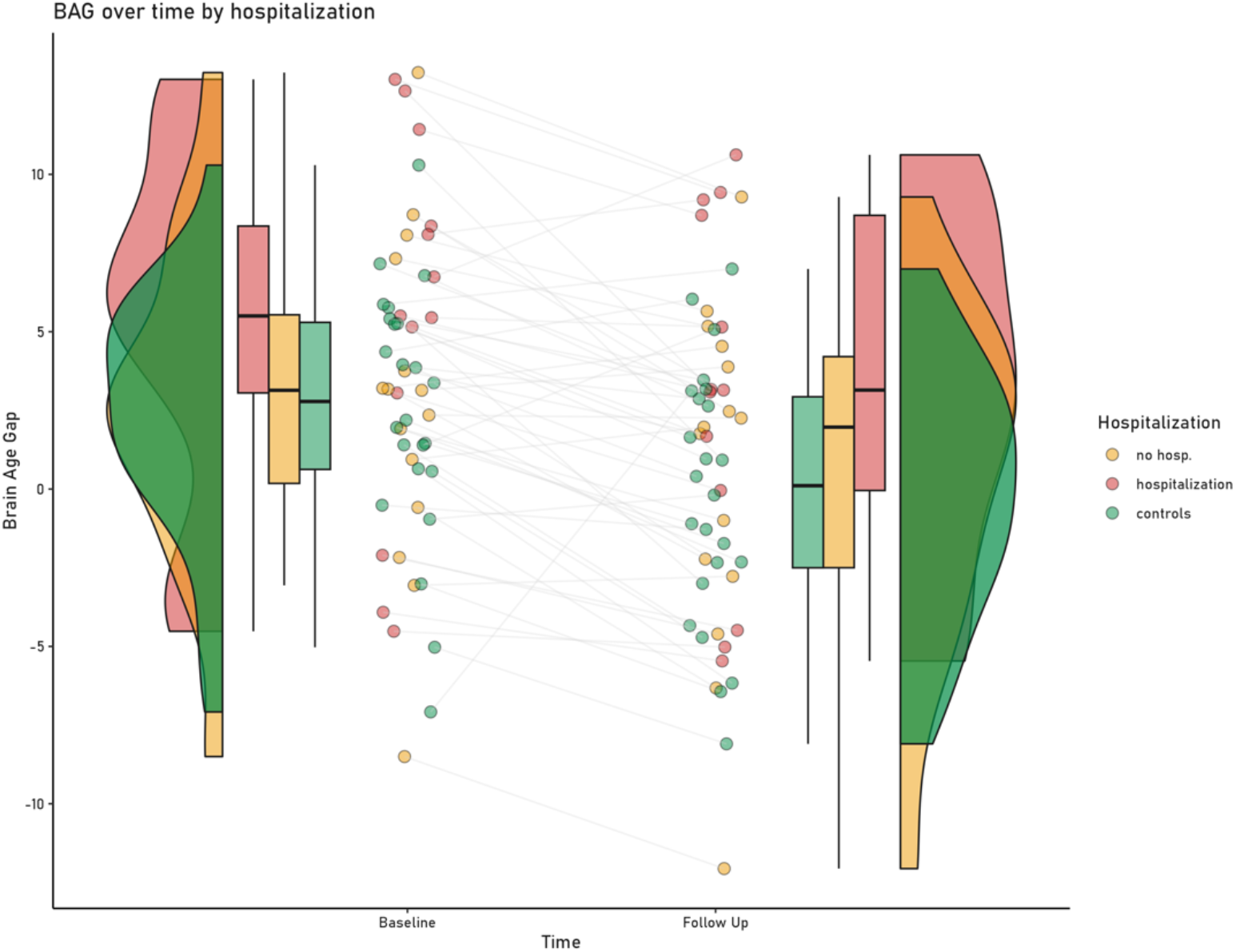
Brain age gap at baseline and follow-up for the healthy control group and patients with or without further hospitalizations between baseline and follow-up time-points.

**Table 1.** Sample characteristics of the Münster-Dublin Longitudinal Cohort.

|  | Healthy Controls (*N* = 37)<br>mean (*SD*) |  | MDD patients (*N* = 32)<br>mean (*SD*) |  | BD patients (*N* = 6)<br>mean (*SD*) |  | Group<br>ANOVA |  |
| --- | --- | --- | --- | --- | --- | --- | --- | --- |
|  | Baseline | Follow Up | Baseline | Follow Up | Baseline | Follow Up | *p*-value | Post hoc |
| Age | 33.73 (12.97) | 42.65 (12.63) | 37.66 (10.62) | 46.23 (9.87) | 34.00 (7.51) | 44.33 (7.06) | .386 | - |
| FU-interval (in months) | / | 107.89<br>(28.55) | / | 104.09<br>(24.13) | / | 126.50<br>(28.31) | .163 | - |
| **Clinical Characteristics** |  |  |  |  |  |  |  |  |
| BDI-I | 2.49 (2.29) | 2.40 (2.59) | 23.50 (11.67) | 10.50 (8.86) | 32.40 (5.68) | 12.50 (8.36) | <.001 | HC < MDD<br>HC < BD |
| HAM-D | / | 1.71 (3.20) | / | 6.34 (7.03) | / | 6.00 (8.88) | .004 | HC < MDD<br>HC < BD |
| YMRS | / | 0.57 (1.04) | / | 0.77 (0.92) | / | 0 (0.00) | .205 | - |
| Number of hospitalizations | / | / | 1.61 (1.46) | 0.91 (1.51) | 3.50 (1.98) | 4.33 (6.95) | .004 | BD > MDD |
| Duration of hospitalizations (in weeks) | / | / | 4.65 (4.11) | 2.18 (4.12) | 15.08 (11.33) | 11.00 (13.91) | .001 | BD > MDD |

*Note.* Hospitalization data, data on patients with bipolar disorder and young mania rating scale are only available for the Münster site. Hamilton depression rating scale and young mania rating scale at baseline were unavailable for a majority of participants and are therefore not reported. *P*-values are reported for ANOVA with subsequent post hoc *t*-tests and between-subjects contrasts of a repeated measurements ANOVA if measurements at Baseline and Follow Up were available.
*Abbreviations:* HC = Healthy Controls, MDD = Major Depressive Disorder, BD = Bipolar Disorder; FU-interval = Follow Up interval; BDI-I = Beck Depression Inventory; HAM-D = Hamilton Depression Rating Scale, YMRS= Young Mania Rating Scale.

## Exploratory Analysis

### Is BAG at baseline predictive of future hospitalizations during the follow-up interval?

The ANOVA model yielded a significant main effect of group on BAG at baseline (F(2,47) = 5.609, p = .001). BAG at baseline was significantly larger in hospitalized than in nonhospitalized patients and controls (t(47) = -3.242, p = .002), while nonhospitalized patients did not differ from controls in their BAG at baseline (t(47) = -0.667, p = .508). Hospitalized patients still differed from controls and nonhospitalized patients when excluding BD patients from the analysis (t(41) = -2.17, p = .035), while nonhospitalized patients did not differ from controls (t(41) =-1.00, p = .323). The difference between hospitalized and nonhospitalized patients also remained significant in a patient-only analysis (t(24) = 2.484, p = .020). BAG at baseline was also associated with the number of hospitalizations during follow-up while controlling for age and inter-scan interval (t(24) = 2.532, p = .018).

Of note, hospitalized and nonhospitalized patients during follow-up did not differ in their hospitalizations before baseline (t(26) = 0.894, p = .379, n.s.).

The logistic regression revealed that BAG at baseline predicted hospitalizations during follow-up (z = 2.103, p = .035). However, the cross-validated mean test accuracy of 64.3% did not reach statistical significance (p = .071).

## Discussion

Using a state-of-the-art brain age prediction model trained on T1-weighted MRI images of 10,000 individuals, we calculated and compared brain age gaps in a sample of 75 patients with affective disorders and healthy controls, measured at two time points with a mean follow-up length of nine years. BAG was higher in patients than in healthy participants. However, while it did not differ between diagnosis (BD and MDD), patients’ disease course was associated with BAG. Patients hospitalized within the follow-up interval showed a higher BAG than healthy controls, while patients without hospitalizations did not differ significantly from healthy participants. Although some studies show no difference in BAG between a healthy population and patients [23, 25, 35], our results are in line with a recent ENIGMA study showing differences between HC and MDD as well as a multi-modal brain age study showing differences between HC and BD [22, 36]. Thus, this provides further evidence for an association of psychopathology with the ageing process of the brain.

In our study, we observe a correlation between age and BAG, with BAG estimates also increasing over time. This can be attributed to the inherent correlation of brain age predictions with chronological age, a characteristic present in both the MCCQR-NN model used here and similar models reported in the literature [13, 30].

There was no evidence of a group-by-time interaction, indicating that the BAG developed similarly within the groups over time, pointing toward a static effect of group (patient status, hospitalization status) at baseline and follow-up. Most importantly, BAG at baseline differed significantly between patients with hospitalizations during follow-up and patients without hospitalizations during follow-up delineating the BAG as a potential biomarker for future chronicity. The exploratory predictive analysis of future hospitalizations by BAG at baseline was not significant. One reason for this could be the limited sample size in this analysis (N = 28 patients were included). Larger studies should follow-up on our descriptive effects using predictive approaches.

In sum, the present study corroborates the idea of considering changes in gray matter in relation to the disease course of affective disorders. Both cross-sectional and longitudinal studies have already established a negative association between gray matter on the one hand and relapse, increased hospitalization, and episodes of illness on the other [27, 28, 37–43]. Our study supports and extends these findings: detrimental disease course manifests not only in local decreases in gray matter, e.g., hippocampus and insula [44, 45], but also in complex parameters based on gray matter measurements, such as BAG.

A higher brain age indicates that patients’ brains appear biologically older than their chronological age, with patients showing a recurrent disease course having a mean BAG at baseline of + 5.7 years. This accelerated brain aging [46, 47] may correspond to alterations of glucocorticoid release or changes in telomere length which have been associated with severity of depression [48, 49]. The early identification of patients with an increased risk for recurrent disease course is key to the development of preventive interventions, including the provision of social psychiatric support measures, the early use of more innovative psychopharmacological therapies, and the use of psychotherapy methods specifically for chronic patients, such as Cognitive Behavioral Analysis System of Psychotherapy [50–54]. However, despite an enormous amount of research, none of the previously identified putative biomarkers have found their way into clinical psychology or psychiatry [55]. Previous structural neuroimaging studies could not reveal that alterations of gray matter may predispose an individual to a recurrent disease course. To tackle this research gap, our longitudinal neuroimaging study, employing a machine learning approach, provides new insights into the relationship between the disease course of affective disorders and gray matter changes. A strength of this study lies in the combination of three innovative methodological aspects. First, it employs a multidimensional measure based on gray matter segments - the BAG. Second, this measure is based on a completely independent brain age model, trained on a substantial and representative German cohort exceeding 10,000 subjects. Third, the study’s longitudinal design explores patients with affective disorders over a large follow-up interval, allowing for a thorough characterization of the patient’s long-term disease course at baseline and during follow-up.

## Limitations

Due to the naturalistic nature of our longitudinal data, no causality can be inferred from our results. However, longitudinal neuroimaging studies are one important way to approach the hen-egg debate of gray matter alterations in affective disorders [38, 39]. Since our results are of correlative nature, we cannot rule out the possibility that not the disease course itself is causally related to the age-related gray matter alterations, but rather that other parameters predispose patients to this outcome, such as genetics, or a history of childhood maltreatment, bullying or increased self-blame [46, 47, 56, 57]). Our sample size was relatively small for our exploratory predictive analysis and the results should therefore be interpreted with caution. While the cross-validated predictive accuracy of 64% significantly exceeds chance levels, its lack of statistical significance suggests a notable variability in the performance estimates across folds. To reliably assess the prognostic biomarker potential of BAG, a larger longitudinal sample will be necessary and should be investigated in future studies.

## Conclusions

The present study revealed the brain age gap as a potential predictor for future chronification in affective disorders. In our analysis, a recurrent course of disease was associated with a larger BAG at baseline, independent of the number of hospitalizations prior to our baseline measurements and medication intake at follow-up. Hence, a higher BAG may predispose patients to a poor outcome and hence could potentially be a relevant predictor of future disease course.

## Data Availability

All raw MRI data in the present study are available upon reasonable request to the authors. All processed data and scripts are available online at https://osf.io/qadxz/.

