## Supplementary Materials for "MRI-derived estimation of biological aging in patients with affective disorders in a 9-year follow-up - a prospective marker of future recurrence"

#### Supplement S1

<u>MRI preprocessing</u>

<u>Münster</u>

T1-weighted high-resolution anatomical images of the head were acquired (Gyroscan Intera 3T, Philips Medical Systems, the Netherlands) at two time points (baseline and follow-up) using a three-dimensional fast gradient echo sequence (turbo field echo), repetition time = 7.4 ms, echo time = 3.4 ms, flip angle = 9°, two signal averages, inversion prepulse every 814.5 ms, acquired over a field of view of 256 mm (feet-head) x 204 mm (anterior-posterior) x 160 mm (right-left), frequency encoding in feet to head direction, phase encoding in anterior-posterior and right-left direction, reconstructed to voxels of 0.5 mm × 0.5 mm × 0.5 mm.

During preprocessing, 3 participants were excluded from further analysis due to movement artefacts at baseline or follow up leading to a final sample of N = 52 (BD = 6, MDD = 22, HC = 24) participants in Münster.

<u>Dublin</u>

During preprocessing, 3 participants were excluded from further analysis due to movement artefacts at baseline or follow up leading to a final sample of N = 23 participants (MDD = 10, HC = 13) in Dublin.

*VBM segmentation and data quality checks*

For VBM data, structural data was processed with the CAT12 toolbox (version 12.6 r1450, http://dbm.neuro.uni-jena.de/cat/) using SPM12 Matlab toolbox with default parameters. In brief, preprocessing steps included segmentation into grey matter, white matter, and cerebrospinal fluid and spatial normalization using the DARTEL algorithm (Ashburner, 2007). Data quality of VBM gray matter segments was verified by the “check homogeneity function” implemented in the CAT12 toolbox and outliers were visually inspected. In consequence, 6 subjects had to be removed from the sample retrospectively, 3 due to excessive head movement, 3 due to inadequate image quality resulting from other artifacts or strong noise. Retest-reliability coefficients for each single subject were calculated by using the covariance structure of the gray matter segments of each individual baseline assessment with its corresponding follow-up image. This procedure was used for additional quality assurance in order to detect outliers as well as to confirm the correct labelling of pre-post image pairs.

#### Sample characteristics for the models using remission vs. hospitalization

##### Table 1. Metric sample characteristics of the patients samples in the remission and hospitalization model

| | **Model with Remission at Follow-Up** | | | | | | **Model with Hospitalization during Follow-Up** | | | | | |
|---|---|---|---|---|---|---|---|---|---|---|---|---|
| | Remitted patients *N* = 23, *M* (*SD*) | | Acute patients *N* = 15, *M* (*SD*) | | Remission ANOVA | | Patients without hospitalization *N* = 15, *M* (*SD*) | | Patients with hospitalization *N* = 13, *M* (*SD*) | | Hospitalization ANOVA | |
| | Baseline | Follow-Up | Baseline | Follow-Up | *p*-value | Post hoc | Baseline | Follow-Up | Baseline | Follow-Up | *p*-value | Post hoc |
| Age | 36.87 (10.68) | 45.83 (10.12) | 37.40 (9.75) | 46.33 (9.29) | .877 | - | 32.40 (8.21) | 42.47 (8.68) | 37.62 (11.57) | 47.31 (10.98) | .189 | - |
| Follow-Up Interval (in months) | / | 107.83 (23.20) | / | 107.33 (27.29) | .953 | - | / | 121.00 (15.00) | / | 117.15 (16.96) | .530 | - |
| **Clinical Characteristics** | | | | | | | | | | | | |
| BDI | 21.10 (11.08) | 5.26 (3.29) | 30.62 (9.57) | 19.08 (7.43) | <.001 | Rem < acute | 16.38 (11.86) | 4.92 (4.34) | 29.00 (7.39) | 15.60 (9.81) | .004 | Non-hosp < hosp |
| HAM-D | / | 2.78 (2.45) | / | 11.67 (8.77) | <.001 | Rem < acute | / | 2.40 (3.29) | / | 10.23 (10.32) | .010 | Non-hsop < hosp |
| YMRS | / | 0.61 (0.85) | / | 0.60 (0.97) | .975 | - | / | 0.73 (0.96) | / | 0.46 (0.78) | .423 | - |
| Number of hospitalizations | 2.11 (1.86) | 2.56 (2.00) | 2.00 (1.49) | 3.50 (5.38) | .089 | - | 1.80 (1.61) | 0 (0) | 2.38 (1.85) | 3.54 (4.63) | .006 | Non-hosp < hosp |
| Duration of hospitalizations (in weeks) | 5.75 (7.51) | 1.64 (3.82) | 8.93 (7.35) | 8.45 (11.28) | .068 | - | 4.133 (2.99) | 0 (0) | 10.06 (9.75) | 8.76 (9.81) | .003 | Non-hosp < hosp |

*Note.* Hospitalization data and young mania rating scale are only available for the Münster site. Hamilton depression rating scale and young mania rating scale were unavailable for many participants at baseline and are therefore not reported. *P*-values are reported for ANOVA with subsequent post hoc *t*-tests and between-subjects contrasts of a repeated measures ANOVA if measurements at Baseline and Follow Up were available.

*Abbreviations:* BDI = Beck Depression Inventory I/II; HAM-D = Hamilton Depression Rating Scale, YMRS= Young Mania Rating Scale.

### Sample characteristics for each site

##### Table 2. Sociodemographic, questionnaire and clinical data of study participants in Münster

| | MDD (N=22) M(SD) | BD (N=6) M(SD) | HC (N=24) M(SD) | P-value according to χ 2-tests or t-tests between clinical groups | P-value according to χ 2-test or ANOVA between all groups |
|---|---|---|---|---|---|
| Age at baseline | 35.1 (10.8) | 34 (7.5) | 29.8 (9.7) | .827 | .205 |
| Age at follow-up | 44.8 (10.7) | 44.3 (7.1) | 40.2 (10.5) | .918 | .297 |
| Interscan interval in months | 117.2 (14.8) | 126.5 (18.3) | 125.2 (18.4) | .207 | .233 |
| Sex (male/female) | 14/8 | 1/5 | 8/16 | .099 | .087 |
| Remitted at follow-up (yes/no) | 15/7 | 3/3 | - | .41 | - |
| hospitalized in interval (yes/no) | 9/13 | 4/2 | - | .262 | - |
| BDI at baseline | 18.9 (11.5) | 32.4 (5.7) | 2.7 (2.4) | .021[a] | <.001[a] |
| BDI at follow-up | 8.8 (9.3) | 12.5 (8.4) | 3.4 (2.6) | .396 | .005[a] |
| HDRS at follow-up | 6.1 (8.3) | 6.0 (8.9) | 2.0 (3.7) | .991 | .107 |
| YMRS at follow-up | 0.8 (0.9) | 0.0 (0) | 0.6 (1) | .053 | .205 |
| Antidepressant medication at baseline (yes/no) | 10/5 | 6/0 | - | .105 | - |
| Antidepressant medication at follow-up (yes/no) | 8/14 | 4/2 | - | .184 | - |

MDD = patients with Major Depressive Disorder; BD= patients with Bipolar Disorder; HC = healthy controls; SD = standard deviation; BDI= Beck Depression Inventory; HDRS = hamilton depression rating scale; YMRS = young mania rating scale. [a]Significant at statistical threshold $P < 0.05$.

#### Table 3. Sociodemographic, questionnaire and clinical data of study participants in Dublin

| | MDD (N=10)<br>M(SD) | HC (N=13)<br>M(SD) | P-value according to χ 2-tests or t-tests between groups |
|---|---|---|---|
| Age at baseline | 43.4 (7.9) | 40.9 (15.5) | .650 |
| Age at follow-up | 49.7 (8.3) | 47.2 (15.3) | .641 |
| Interscan interval in months | 75.2 (12) | 76 (9.9) | .862 |
| Sex (male/female) | 3/7 | 4/9 | .968 |
| Remitted at follow-up (yes/no) | 5/5 | - | - |
| BDI at baseline | 31.7 (6.6) | 2.1 (2.1) | <.001[a] |
| BDI at follow-up | 13.3 (7.8) | 0.4 (0.7) | <.001[a] |
| HDRS at follow-up | 7 (2.6) | 1.3 (2.1) | <.001[a] |
| Antidepressant medication at baseline (yes/no) | 9/1 | - | - |
| Antidepressant medication at follow-up (yes/no) | 1/12 | - | - |

MDD = patients with Major Depressive Disorder; BD= patients with Bipolar Disorder; HC = healthy controls; SD = standard deviation; BDI= Beck Depression Inventory; HDRS = hamilton depression rating scale.
[a]significant at statistical threshold $p < .05$.
